# Predicting long term clinical outcomes in Parkinson’s Disease using short term rating scales

**DOI:** 10.64898/2026.03.27.26349548

**Authors:** Matthew Burnell, Cristina Gonzalez-Robles, Marie-Louise Zeissler, Michèle Bartlett, Caroline S Clarke, Carl Counsell, Michele T. Hu, Tom Foltynie, Camille B Carroll, Michael Lawton, James Carpenter, Yoav Ben-Shlomo, the EJS-ACT PD Consortium

**Author notes:** **Corresponding Author:** Matthew Burnell, MRC CTU 90 High Holborn 2nd Floor, London, WC1V 6LJ, United Kingdom; 0044 7868 163181.

## Abstract

**Background:** Most trials of Parkinson’s disease (PD) measure progression over a short to medium time-period using continuous rating scales that may be hard to interpret and less meaningful for patients. There is a lack of evidence connecting changes in these scales to changes in outcomes important to patients.

**Objectives:** We present causal modelling to translate the causal, short-term disease-modifying treatment effects on functional rating scales to the 10-year risk of serious clinical progression milestones.

**Methods:** We selected four important clinical milestones of disease progression from the Oxford Parkinson’s Disease Centre “Discovery” cohort: dementia, any falls, frequent falls, and mortality. We proposed a causal framework for our research objectives so we could model the potential impact of a 30% reduction in disease progression slopes (“treatment effect”) using the summation of parts I and II of the Movement Disorders Society Unified Parkinson’s Disease Rating Scale (*UPDRS12)*. This outcome was regressed on time to milestone using flexible parametric survival models. Marginal predictions of survival and survival difference at year 10 were then calculated for the Discovery cohort, and a counterfactual cohort applying the treatment effect to estimate the relative and absolute reductions for the four clinical milestones.

**Results:** The model increase in risk for each unit change in the *UPDRS12* were as follows: dementia hazard ratio (HR)=1.52 (95% Confidence Interval (CI) 1.36-1.70), any falls HR=1.37 (95% CI 1.29-1.46), frequent falls HR=1.68 (95% CI 1.49-1.89), mortality=1.29 (95% CI 1.17-1.42). These models led to marginal predictions of absolute reductions, when the progression was reduced by 30%, between 4.0% (mortality) and 7.5% (frequent falls) at 10 years follow up.

**Conclusions:** We have demonstrated how a treatment effect in a trial specified in terms of a progression change of a rating scale can be contextualised into a long-term reduction in the probability of clinically relevant milestones. Whilst we have used PD as our exemplar, we believe this methodological approach is generalisable to other chronic progressive diseases where trials are often limited to a relatively short follow-up period and use some scalar measure of progression, but significant clinical milestones usually take longer to be observed.

**WHAT IS NEW?:** *Key findings:* A clinically significant reduction in Parkinson’s progression, as targeted in a major new multi-arm multi-stage trial, could result in absolute reductions for key “milestone” events dementia, falls and mortality of between 4-7.5% after 10 years

*What this adds to what is known?:* We have shown how one could model the relationship between changes to a short-term measure of disease progression and significant long-term outcomes (“milestones”) providing meaningful context for participants, triallists and funders alike concerning the treatment effect of a disease-modifying therapy (DMT).

*What is the implication and what should change now?:* This method could be applied to the DMT trialled for any chronic progressive diseases where the important clinical outcomes are typically not observed within the trial’s duration

## 1. INTRODUCTION

Parkinson’s disease (PD) is a slowly progressing neurodegenerative condition encompassing motor and non-motor signs and symptoms[1-3] with a significant physical, emotional and financial burden to patients, care partners, and society. There is an urgent need for effective disease-modifying therapies (DMTs)[4, 5] to slow the progression trajectory which in the long-term should lead to better outcomes for patients, by delaying the severity and impact of the disease. However, PD progresses slowly, is heterogeneous[6-11] and symptomatic therapy may mask motor disease progression on clinical scales[12, 13]. Therefore, trials of between 2 to 5 years are recommended by regulators to capture true disease-modifying benefits[14]. Most trials, however, are either shorter than this or fall within the lower range of this time frame[15, 16] ignoring the much longer life expectancy (14+ years depending on age) of these patients[17]. Accurately projecting from short-term outcomes to the long-term benefits of new disease-modifying treatments (DMTs) is crucial to communicate effects to patients and clinicians. Such milestones are directly relevant to patients and have greater health-economic consequences than short-term scale changes[18]. It also equips policy makers to evaluate cost-effectiveness and suitability of novel DMTs for their potential integration into routine standard of care. For example, in the UK, the multiple sclerosis risk sharing scheme evaluated treatments using observational data to predict clinical benefits at 10 years as none of the original trials went beyond five years[18].

The current study is motivated by the Edmond J Safra Accelerating Clinical Trials in PD (EJS ACT-PD) initiative, which is developing a multi-arm multi-stage (MAMS) PD trial platform trial[19]. EJS ACT-PD is designed to detect the causal effect of DMTs on the MDS-UPDRS scale over 3 years; we wish to complement this with an estimation of the causal effect on more meaningful disease milestones over 10 years. We believe our practical approach will be helpful not just for PD trials but also for trials of other slowly progressive chronic disorders.

## 2. METHODS

### 2.1 Cohort details

Our data come from the Oxford Parkinson’s Disease Centre (OPDC) “Discovery” cohort. This recruited people with Parkinson’s (PwP) within 3.5 years of diagnosis (“early” PwP) from the Thames Valley Region (between 2010-2016)[20]. Participants had a clinical diagnosis of idiopathic PD diagnosed according to the UK PD Brain Bank criteria by a neurologist or geriatrician with a specialist interest in PD; the validity of diagnosis was additionally checked by the study neurologist at screening. Exclusion criteria included non-idiopathic parkinsonism, dementia preceding PD by one year suggestive of dementia with Lewy bodies, and cognitive impairment precluding informed consent[21]. Consenting participants were assessed every 18 months at a research clinic (or by telephone or videoconference during COVID-19).

### 2.2 Definition of short-term predictor and long-term milestones

We have used the summation of parts I and II of the MDS-UPDRS clinical scale, as supported by the original developers of the MDS-UPDRS[22], as our short-term predictor of disease progression. This is mostly based on patients’ report of motor and non-motor experiences of daily living and is the primary outcome of the planned EJS ACT-PD trial[23] based on extensive input from a Patient and Public Involvement and Engagement (PPIE) panel (see[23] for detailed explanation). Our predictor variable (rate of disease progression) was an individual’s MDS-UPDRS slope derived from their longitudinal MDS-UPDRS parts I and II sum scores (“*UPDRS12*” slope) across visits 1 to 3 (approximately every 18 months). The follow-up period of three years reflects the trial duration in EJS ACT-PD. Our selected long-term clinically relevant outcomes (milestones) were ranked amongst the top five research priorities for PD[29]: self-reported history of any falls, frequent falls, dementia and all-cause (by necessity) mortality. Supplement 1 gives detailed definitions.

### 2.3 Statistical methods

#### 2.3.1 Assumptions

Because the data come from an observational study, the relationship between early-disease changes in MDS-UPDRS and incidence of long-term clinical milestones may be confounded. Figure 1 is a directed acyclic graph (DAG) showing the direct and indirect effect of a DMT on long-term clinical milestones which may be partially mediated by its effect on MDS-UPDRS progression. If we had data from a randomised controlled trial we could estimate both, giving the total effect. However, with our data we can only estimate relationships to the right of the dashed line.

**Figure 1.**
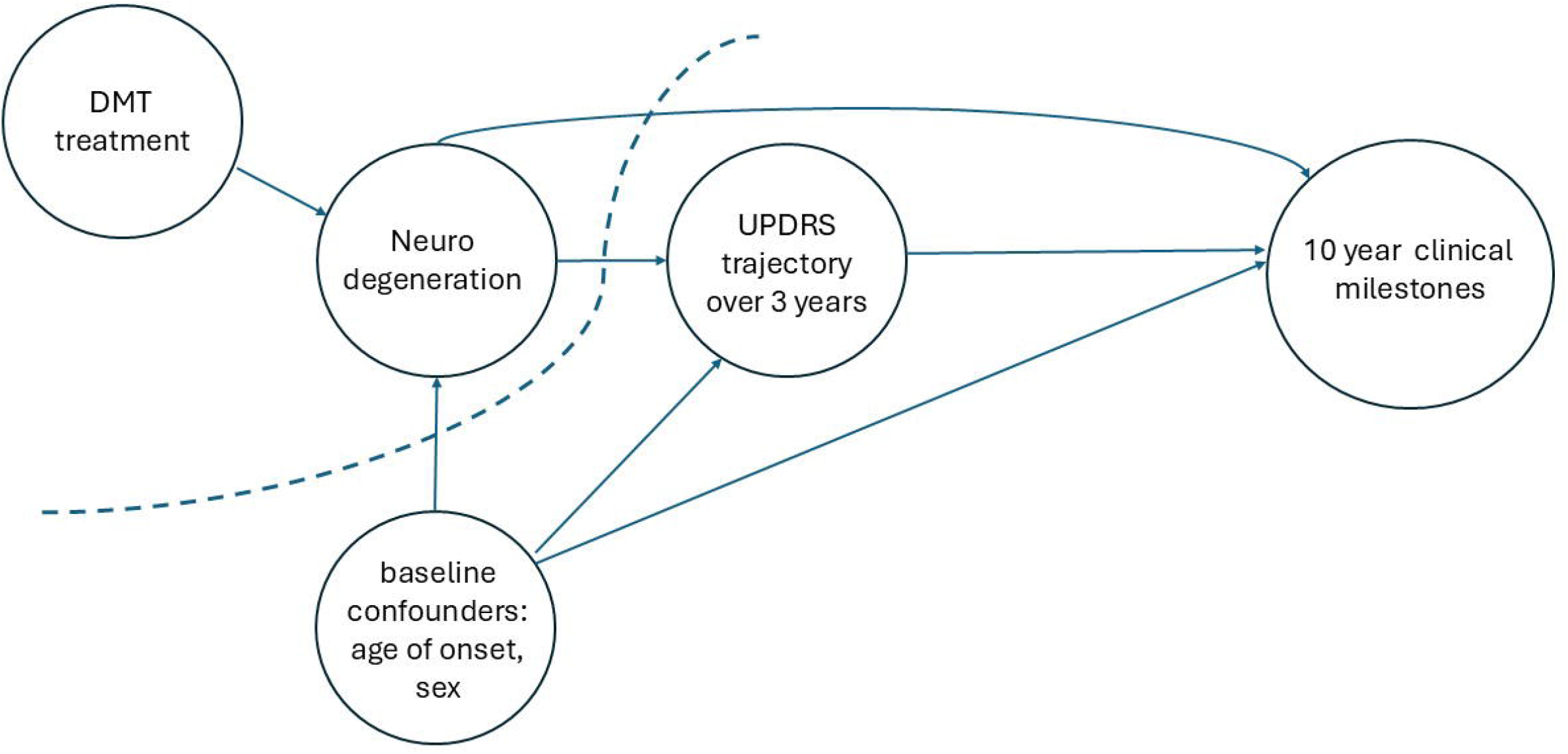
Directed acyclic graph Directed acyclic graph (DAG) showing the direct and indirect effects of a DMT on long-term clinical milestones

To use the results of our analyses to estimate the causal long-term effect of DMTs on incidence of clinical milestones, we have made the following assumptions:

1. The relevant milestones are deaths, falls and dementia.
2. The principal confounders of the causal effect of MDS-UPDRS trajectory are sex, age at baseline and time since diagnosis with no important unmeasured confounders.
3. There was no clinically meaningful measurement error.
4. There was no effect modification of treatment by age, time since diagnosis and sex
5. Within each stratum of the confounders, we have a range of MDS-UPDRS trajectories (“positivity”).
6. There is a causal effect of disease modification (change in MDS-UPDRS) on milestones, so if a trial shows a DMT reducing the rate of increase in MDS-UPDRS, then long-term use will delay the clinical milestones. This is plausible because MDS-UPDRS measures the phenotypes forming the clinical milestones.
7. The effect of treatment on the trajectory persists for 10 years. This is plausible if a DMT is well tolerated and is taken indefinitely.
8. Parametric models are acceptable.

#### 2.3.2 Individualised slopes

We derived short-term progression by regressing *UPDRS12* score on study time from first visit (*t*=0) using a mixed model with correlated random intercept and slope parameters. Time was treated linearly in the model following inspection of *UPDRS12* scores over time including a locally weighted smoother (Supplement 2). Individualised slopes were created by summing the overall mean slope to each subject’s random slope estimate, which effectively ‘borrow’ from the complete dataset and shrink the “Best Linear Unbiased Predictions” (BLUP) towards the overall mean slope. This makes them more robust than a simple ‘best fit’ estimate, especially when the number of data points per person is small[24]. Assuming “missing at random”, predictions can still be made for those with only 1 or 2 available data points[24].

#### 2.3.3 Survival modelling

Survival analysis measured the strength of association between (BLUP) based slopes (“exposure”) with the long-term outcomes using flexible parametric models[25] that fit restricted cubic splines in the log-cumulative hazard scale using 2 or 3 degrees of freedom (DF). These models allow for an arbitrarily close fit to the underlying hazard function and provide an easy way to model any non-proportionality in the hazard ratio. Analysis time was defined as time from baseline visit to the outcomes specified above or censoring at last available visit. As dementia and falls outcomes were ascertained at visits, technically these events were interval censored. Models were checked for non-proportionality by investigating the improvement in fit for models adding a time by predictor interaction. We adjusted for sex (assigned at birth), age at baseline visit and time since diagnosis as plausible confounders of the short-term predictor and longer-term outcomes. Any outcomes already recorded at the baseline visit (*t*=0) were necessarily excluded from the models by their null contribution to the likelihood function.

#### 2.3.4 Marginal predictions

Relevant hazard and marginal survival functions were calculated post-estimation and plotted. We derived and plotted the survival difference for a hypothetical 10-year trial comparing the PD progression as observed from the Discovery cohort (‘untreated’) versus a counterfactual (‘treated’) cohort by applying a 30% reduction in progression to the ‘untreated’ cohort, as the EJS ACT-PD trial protocol considered this a clinically meaningful benefit. All survival predictions were marginalised over the distribution of individuals’ covariate values from the sample. If an individual’s slope estimate was zero or negative (‘improving’) then the slope value used for the contrast ‘treated’ cohort was left unchanged vis-a-vis the ‘untreated’ Discovery cohort. In these cases, it did not make sense for them to have a ‘30% improvement’.

We repeated the marginal predictions for a cohort with a mean age of 60 (but identical standard deviation) to examine the clinical meaningfulness of a 30% treatment effect reduction in a younger PD trial population, given the mean age of participants is typically younger (62 years) in trials than in in the Discovery cohort (67.9 years)[15].

#### 2.3.5 Sensitivity analyses

There is a potential risk of reverse causality, given the time-period covering progression slope overlaps with the first 3 years of the survival analysis e.g. dementia onset could contribute to faster progression in the first 3 years. We performed two sensitivity analyses to examine this: (i) we only included events after the first 3 years of observation (ii) we repeated our models restricted to subjects whose baseline visit was within the first 18 months of disease diagnosis as these outcomes are rare unless they have another parkinsonian condition.

All analyses were performed using Stata 17.0 (StataCorp 2021) using user-written packages `stpm2` and `standsurv`[26, 27]. Stata code used to perform the analysis is provided in Supplement 3.

## 3.0 RESULTS

More detailed results are provided in Supplement 1.

### 3.1 Basic descriptives

We analysed 3667 observations from 958 participants (up until 17/05/2022) with 77% having at least 4 visits and 33% at least 6 visits. The median age at first visit was 67.9 years (IQR: 61.4-74.1; range: 32.2-90.5) with a male predominance (63.9%). The mean time since clinical diagnosis at baseline was 1.25 years (median=0.96; IQR=0.44-1.98; maximum=3.49 years).

The overall mean slope for *UPDRS12* was 2.08 points increase per year (standard error [se]=0.11, median slope 1.96, intercept 17.5 (se=0.33)). Two subjects were excluded from all models as they had no *UPDRS12* data in the first 3 years. Furthermore, 42 patients (4.4%) had a slope less than 0, so that their slope was left unchanged when included within the ‘treated’ cohort. The mean slope for those who died and those who survived were 2.5 and 2.0, both before and after 3 years, suggesting that missing visits in years 0-3 do not induce obvious bias in *UPDRS12* slope estimates (Supplement 4).

### 3.2 Survival modelling and predictions

The results for a unit increase in *UPDRS12* slope for all four outcomes both in relative and absolute terms are shown in Table 1. We have also graphically shown how the hazard rates, marginal survival probabilities and marginal survival differences vary with follow-up period (see Figures 2 to 4 respectively). More details about the number of subjects in the analysis, events, exclusions and follow-up are provided in Supplement 1. Supplement 5 collates complete prediction results for all outcomes assuming a 30% and, additionally, 50% reduction in progression rate.

**Figure 2.**
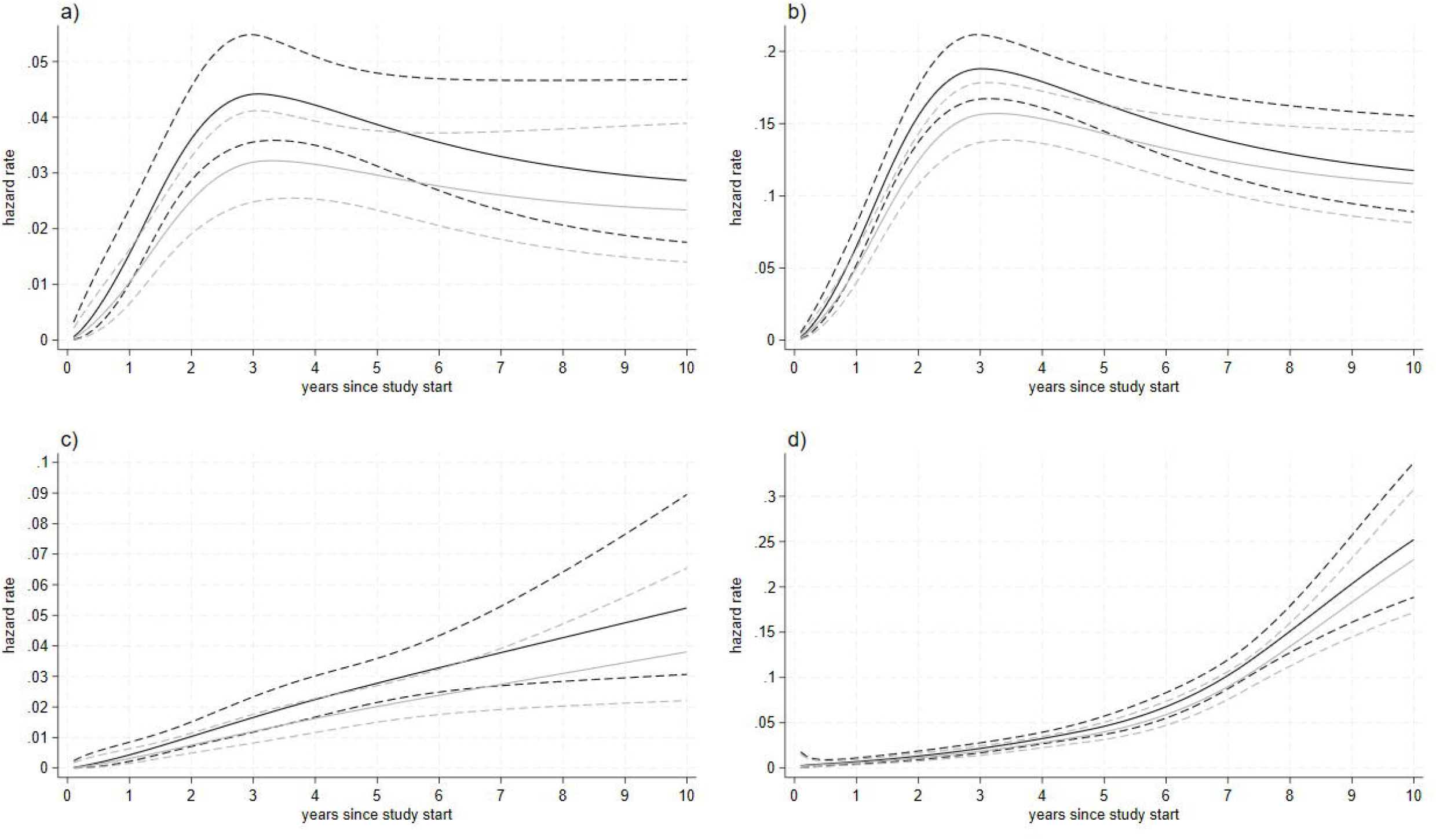
Marginal hazard functions Marginal hazard functions (solid line) with 95% confidence bands (dashed lines) over a 10-year range for the following clinical milestones: a) dementia b) any falls c) frequent falls d) all-cause mortality, contrasting the Discovery ‘untreated’ (dark) and ‘treated’ cohort assuming a 30% reduction in progression (light).

**Figure 3.**
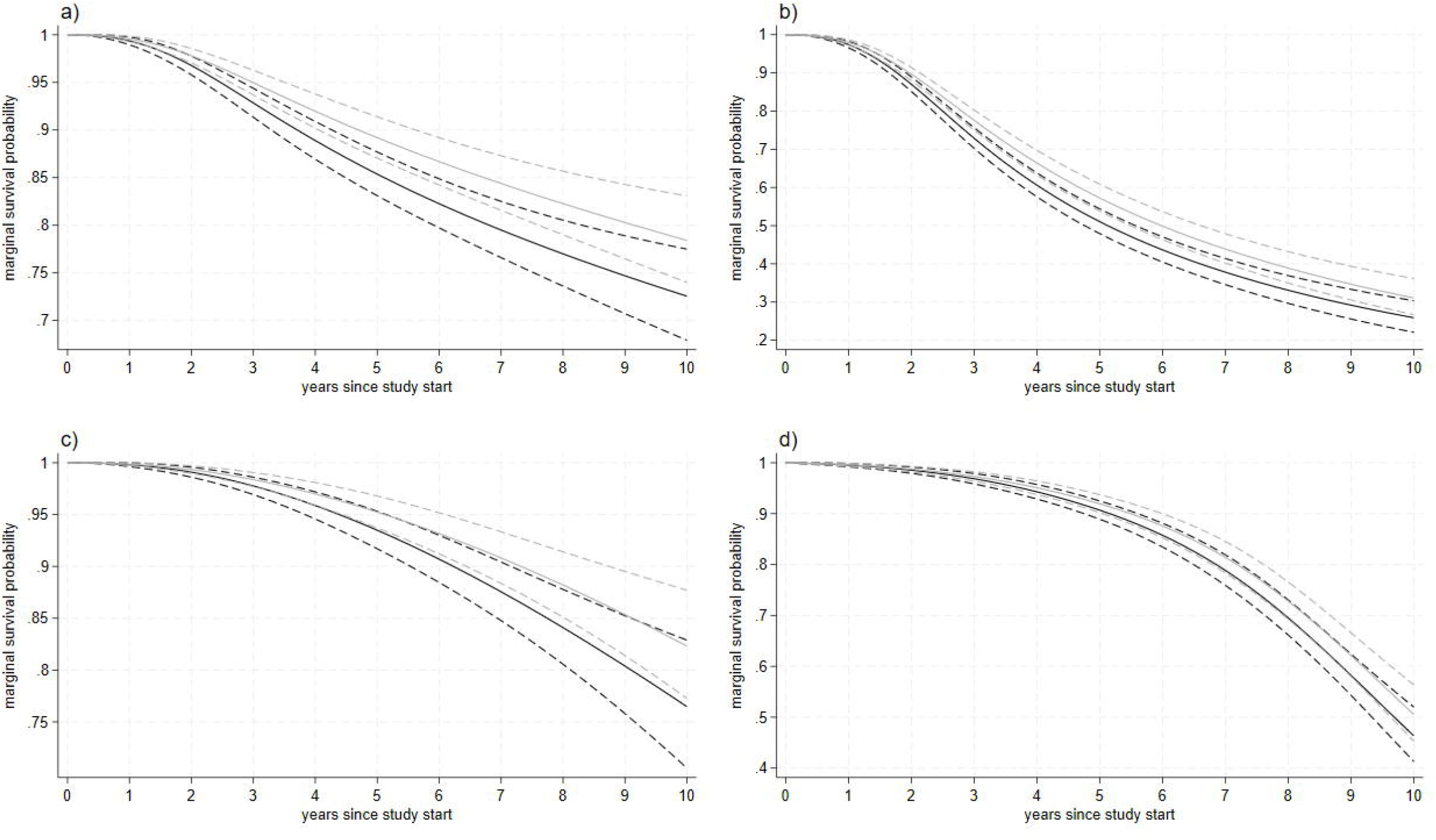
Marginal survival functions Marginal survival functions (solid line) with 95% confidence bands (dashed lines) over a 10-year range for the following clinical milestones a) dementia b) any falls c) frequent falls d) all-cause mortality, contrasting the Discovery ‘untreated’ (dark) and ‘treated’ cohort assuming a 30% reduction in progression (light).

**Figure 4.**
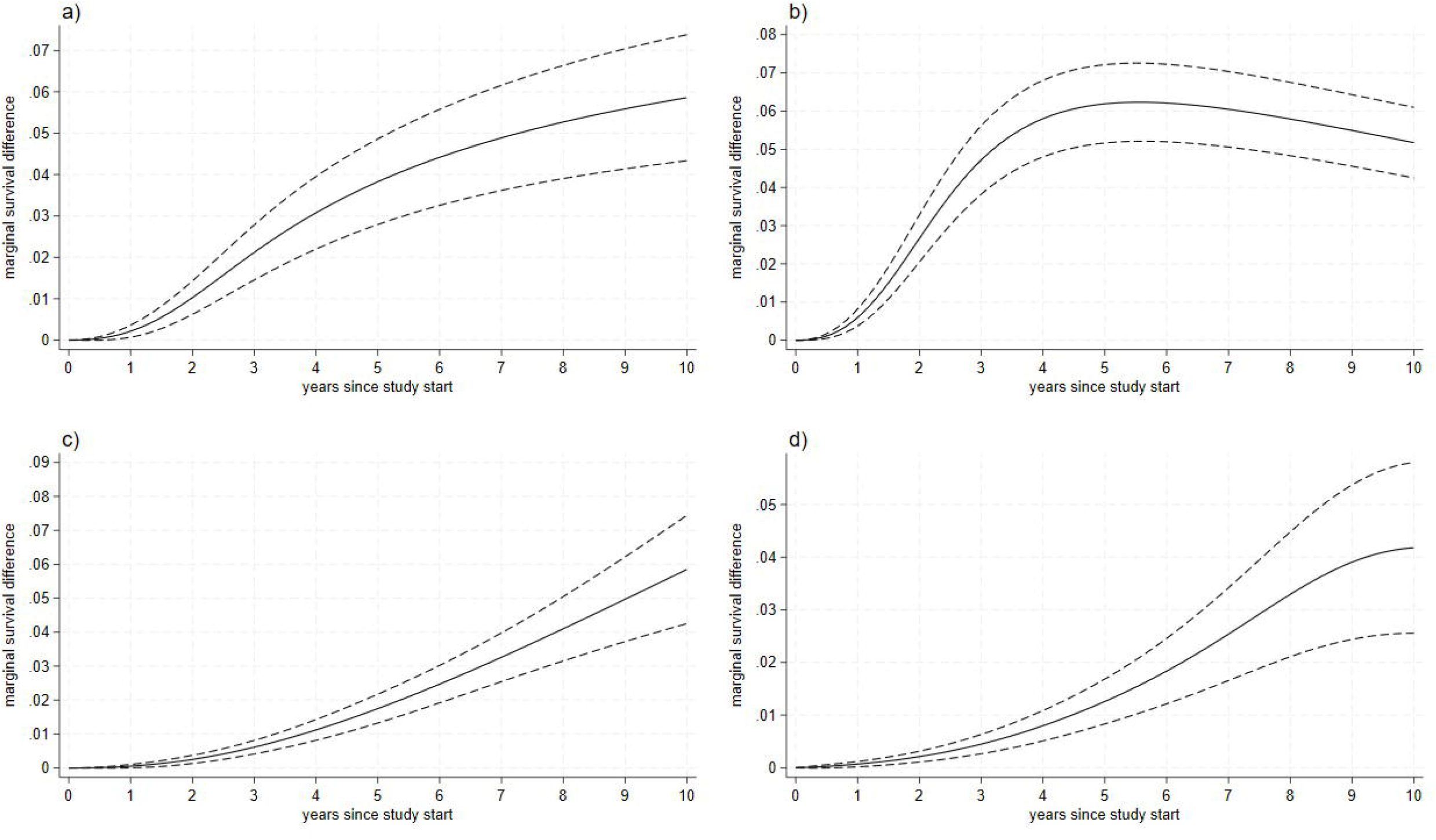
Marginal survival difference functions Marginal survival difference functions (solid line) with 95% confidence bands (dashed lines) over a 10-year range for the following clinical milestones a) dementia b) any falls c) frequent falls d) all-cause mortality, presenting the absolute risk difference between the Discovery ‘untreated’ and ‘treated’ cohort assuming a 30% reduction in progression.

**Table 1.**
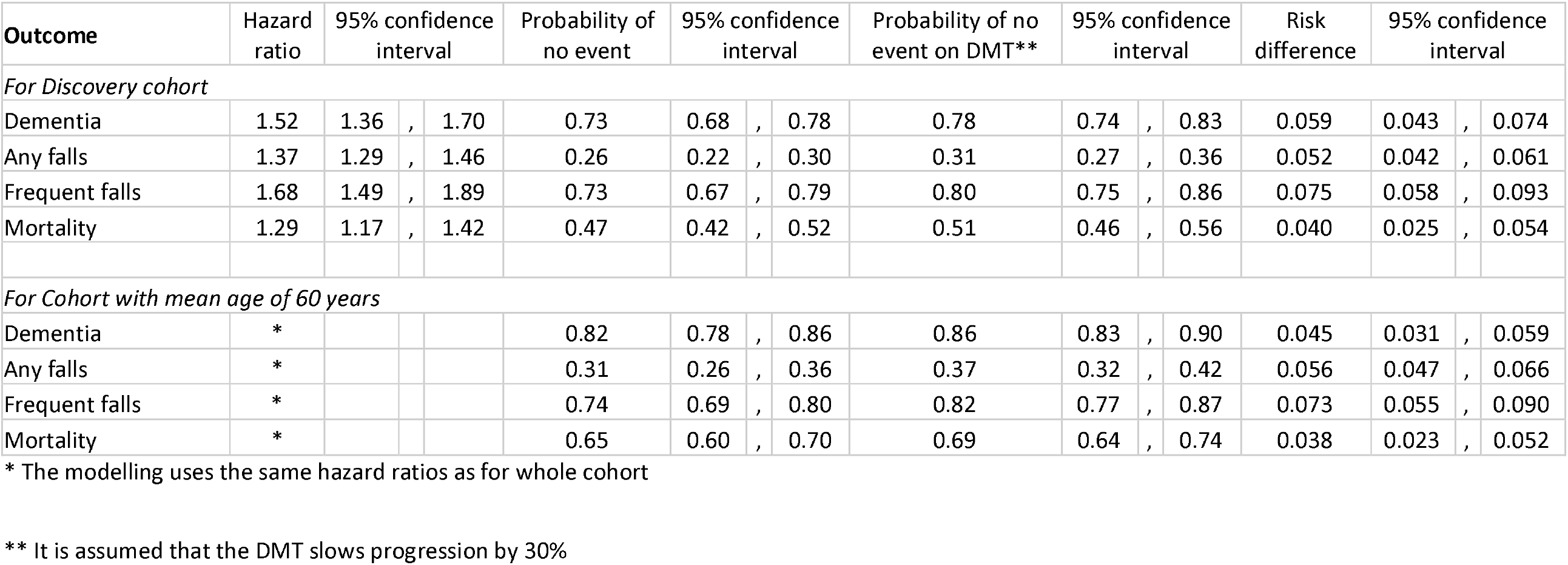
Table of hazard ratios for *UPDRS12* when modelling dementia, any falls, frequent falls and mortality; plus year-10 marginal survival and survival difference estimates for the Discovery ‘untreated’ and ‘treated’ cohort. Lower portion contains equivalent survival estimates for younger cohorts (mean age=60).

A unit increase in the *UPDRS12* slope was associated with an estimated hazard ratio (HR) of 1.52 (95% Confidence Interval [CI] 1.36-1.70; p<0.0001) for dementia. There is a large increase in the marginal hazard over the first 3 years because of the exclusion of any cases at baseline (Figure 2a). After 3-4 years, the hazard declines gently over the remaining time-period but with widening uncertainty. The DMT benefit in absolute terms at 10 years was a predicted 5.9% (95% CI: 4.3-7.4%) reduced risk of a dementia outcome (number needed to treat to benefit (NNTB) = 17).

The estimated HR per 1-unit UPDRS12 slope increase for any falls was 1.37 (95% CI 1.29-1.46; p<0.0001) and 1.68 (95%CI 1.49-1.89; p<0.0001) for frequent falls. The shape of the (baseline) hazard function for any falls was similar to that of dementia, but with greater absolute values given this outcome is more common, whilst the shape was different for frequent falls, increasing monotonically. The projected difference in survival probabilities at 10 years was 5.2% (95%CI: 4.2-6.1% and NNTB=20) and 7.5% (95%CI: 5.8-9.3% and NNTB=13.3) for any and frequent falls, respectively.

The mortality hazard ratio per 1-unit UPDRS12 slope increase was 1.29 (95% CI 1.17-1.42; p<0.0001) and the hazard function accelerated quickly, especially after year 6. The survival difference was predicted to be a 4.0% reduction (95%CI: 2.5-5.4% and NNTB=25.0). For all models, there was no evidence of non-proportional hazard functions associated with *UPDRS12*.

We repeated the 10-year survival predictions for a population with a mean age of 60 (7.3 years younger). This younger cohort had higher survival predictions (Table 2b and Supplements 6 and 7) but similar survival differences.

**Table 2.**
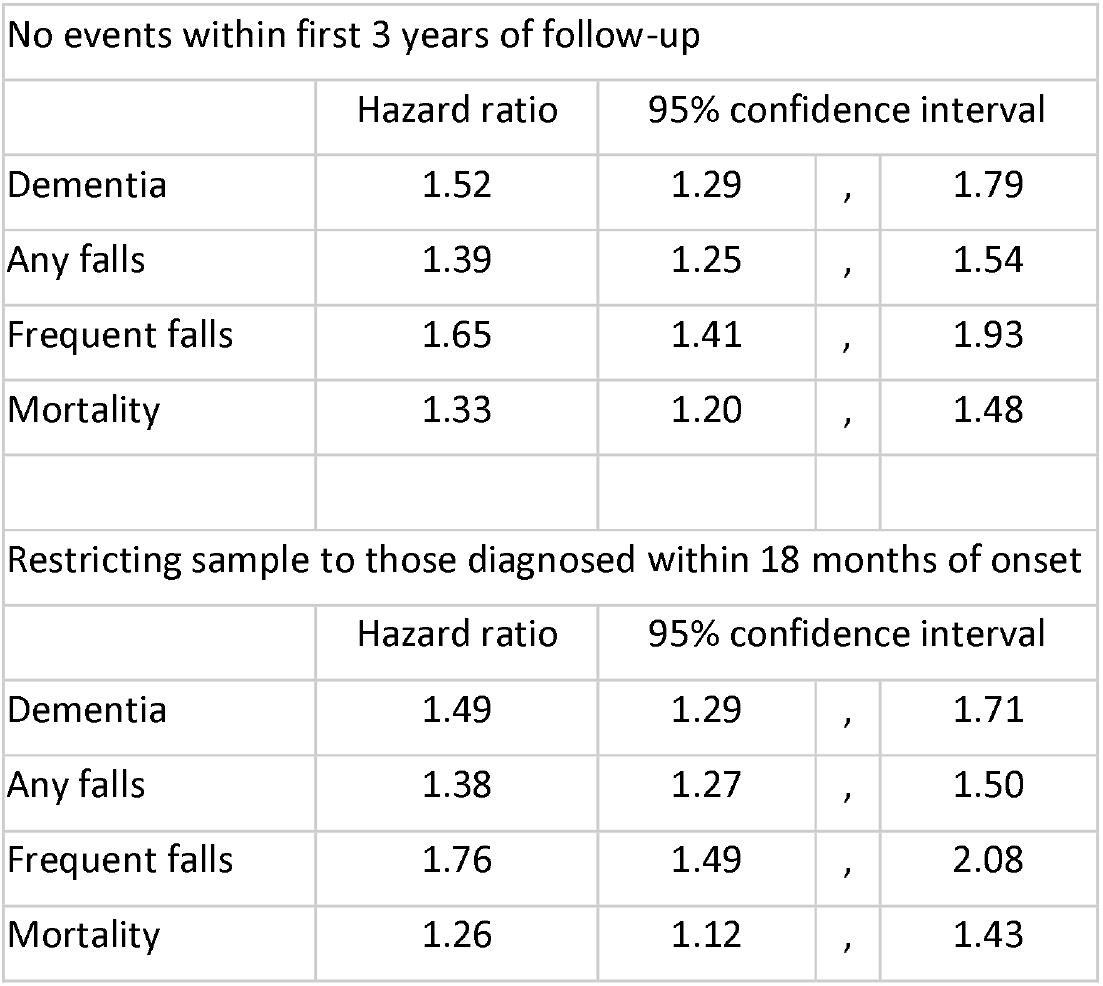
Sensitivity analysis table of hazard ratios for *UPDRS12* when modelling milestones dementia, any falls, frequent falls and mortality. A) excludes events from 3 years and B) restricts analysis to participants diagnosed within 18 months of study start.

Table 2 shows the sensitivity model results for (a) those with an event of interest only after the first 3 years and (b) those whose time since diagnosis was less than 18 months. The point estimates only changed slightly, though confidence intervals were wider than those using the whole cohort.

## 4.0 Discussion

Few PD trials are conducted for longer than 6 years because of (i) cost (ii) participant attrition (iii) potential treatment contamination and (iv) ethical considerations due to the need to maintain blinding and equipoise if related results are published. Evidence of drug benefit is ascertained over a relatively short period (2-3 years) where meaningful milestones are rare, and this is especially true for RCTs of novel and/or costly therapies. PD trials of DMTs traditionally recruit early onset or even prodromal participants, as it is assumed that interventions may be more effective at this stage. Whilst clinicians and PwP may intuitively feel that a 30% improvement in motor and non-motor progression must be clinically relevant, evidence of “real-world” impact would allow for more informed discussions and better speak to policymakers, payers and participants.

We have attempted to overcome these limitations using a large, high-quality dataset of people with early PD with regular long-term follow-up. It is important to note we are not attempting to validate a surrogate outcome, which typically have modest correlations with clinically important outcomes[28], but calibrate a rating scale already judged as relevant for use in PD trials[22]. We modelled the potential impact of any future DMT that reduced the trajectory of parts I and II of the MDS-UPDRS scores on relative and absolute risk reductions of meaningful milestones over 10-years. The choice of a 30% reduction was based on the detailed consideration of a multi-arm multi-stage (MAMS) trial platform developed by the EJS ACT-PD consortium[19]. Our models are essentially “proof of concept”, and the methods we have outlined could generate disease-specific predictions for different estimates of drug benefit (say, from 20 to 50% relative decline in primary outcome), varying follow-up durations (say, 5 to 15 years) and the relevant milestones.

Our results show that an annual unit increase in a combined MDS-UPDRS parts I and II score is associated with an increased hazard of between 30% to 70% across all our outcomes, with rare outcomes showing greater hazard ratios (dementia, frequent falls). If a novel therapy can reduce the rate of increase of MDS-UPDRS parts I and II scores by 30%, this may translate to an absolute reduction of between 4% to 7% in the frequency of these meaningful outcomes - or an NNTB of between 15 to 25 patients - at 10 years. The absolute benefits will be larger over a longer follow-up period. The NNTB numbers were only slightly larger when applied to younger populations with lower absolute risk. For comparison, the 10-year NNTB under the UK National Institute for Health and Care Excellence (NICE) guidelines for statins in individuals at high versus low risk of developing heart disease (20% and 5% risk) is 14 to 50[30].

There are several strengths to this analysis. We have used a relatively large, contemporary, high quality and still active dataset that recruited PwP early in their disease course. Diagnosis was made by specialist neurologists excluding subjects with atypical parkinsonism, with a worse prognosis. Disease progression was tracked through specialist research clinics, phone or video clinics every 18 months. Attrition due to dementia was captured as part of the exit protocol. We used sophisticated and flexible statistical models, examined for reverse causation and generated predictions for younger populations more typical of RCTs[15].

The following limitations need consideration. (i) Our modelling assumes that a 30% treatment reduction in the MDS-UPDRS trajectory remains constant up to 10 years if patients continue to receive long-term treatment, unless there are adverse effects. If the therapeutic effect wanes over time, then our models will have overestimated the treatment benefit. (ii) This analysis does not replace a full health-economics evaluation. We did not include other outcomes such as quality-of-life and service-use which will also contribute to any decision about cost-effectiveness. (iii) We had to use all-cause mortality rather than PD-specific mortality. This likely resulted in an under-estimation of the PD mortality benefits assuming the treatment has no effect on non-PD deaths (e.g. through side effects or comorbidities). (iv) If subjects who die are more likely to develop PD dementia, this will induce informative censoring for that milestone, though methods for competing risks could be used instead[31]. (v) Our cohort represents PwP from South-East England, under-representing PwP who are older and from minority ethnic groups[21]. Our results may or may not be generalizable to other populations, so replication is required. (vi) Subjects with only baseline data were excluded from the survival analysis which could have introduced selection bias if there was informative censoring. (vii) We assumed *UPDRS12* data was missing-at-random when deriving individual slope estimates. It is possible that those with incomplete data missed visits due to poorer health and hence their slope estimates could be biased downwards. Pattern-mixture modelling could be used if this were a concern, although such methods suggested dropout is not influential on trajectory estimation for an earlier realisation of this cohort[32, 33].

We believe our methodology can be applied to other progressive chronic diseases to translate short-term benefits on rating scores to long-term clinical milestones. This is helpful for future RCT design and could be incorporated in sample size considerations. Ultimately, whether these reductions in 10-year outcomes are sufficiently valuable to support adopting a new treatment, in addition to the short-term impact on motor and non-motor activities of daily living, needs to be assessed in balance with cost-effectiveness, side effects, patient and clinician-acceptability. We hope that the approach illustrated in this research will help inform decision-making for other researchers, as well as helping the medical and lay community understand the longer-term impact of short-term trials that try to alter the natural history of PD.

## Data Availability

Data from the Oxford Parkinson's Disease Centre Discovery cohort is available on request from supplied URL. Analysis code is available as a Supplement.

https://www.dpag.ox.ac.uk/opdc/research/external-collaborations

## Financial disclosure related to the manuscript

M.Bu. has received research support from the Edmond J. Safra (EJS) Foundation (EJS Accelerating Clinical Trials in Parkinson’s Disease [EJS ACT-PD] project) and is supported by Medical Research Council grants MC_UU_00004/06 and MC_UU_00004/09.

C.G.R. has received research support from the Edmond J. Safra (EJS) Foundation (EJS Accelerating Clinical Trials in Parkinson’s Disease [EJS ACT-PD] project) and has no conflict of interest to report.

M.-L.Z. has received research support from the Edmond J. Safra (EJS) Foundation (EJS Accelerating Clinical Trials in Parkinson’s Disease [EJS ACT-PD] project) and has no conflict of interest to report.

M.B. has no financial disclosures relating to the content of this article and has no conflict of interest to report.

C.S.C. is supported by the National Institute for Health and Care Research University College London Hospitals Biomedical Research Centre.

C.C. has no financial disclosures relating to the content of this article and has no conflict of interest to report.

M.T.H. has received funding from Parkinson’s UK for the Discovery cohort (grant reference J-1403) and has no conflict of interest to report.

T.F. is co-chief investigator for the EJS ACT-PD initiative and co-chief investigator for the EJS ACT-PD MAMS platform trial and has received research funding from Edmond J Safra Foundation.

C.B.C. is co-chief investigator for the EJS ACT-PD initiative and co-chief investigator for the EJS ACT-PD MAMS platform trial and has received research funding from Edmond J Safra Foundation.

M.L. received fees for advising on a secondary analysis of an RCT sponsored by North Bristol NHS trust.

J.C. is supported by Medical Research Council grants MC_UU_00004/06 and MC_UU_00004/09.

Y.B.S. has no financial disclosures relating to the content of this article and has no conflict of interest to report.

## Ethics approval statement

The Oxford Parkinson’s Disease Centre Discovery cohort was approved by NRES Committee, South Central Oxford A Research Ethics Committee, Reference number 16/SC/0108

## Data sharing

Data from the Oxford Parkinson’s Disease Centre Discovery cohort is available on request from https://www.dpag.ox.ac.uk/opdc/research/external-collaborations. Analysis code is available as a Supplement.

## AI Statement

No use AI was made anywhere in this submission

## Acknowledgments

We sincerely thank the PPIE members in EJS ACT-PD for their invaluable contribution to this work. We thank Prof. Barker, co-chair of the EJS ACT-PD working group for his critical review of the manuscript.

## Authors’ Roles

Design: Y.B.S, J.C., M.Bu., M.L., Upholding patient perspective in design, and interpretation of results: M.B.; Data collection and management: M.T.H., Y.B.S., M.L.; Analysis: M.Bu., Writing: M.Bu., C.G.R., M.-L. Z, Y.B.S, J.C., Editing of final version of the manuscript: Y.B.S, J.C., M.L., M.T.H, M.-L. Z, C.S.C., M.B., C.G.R., M.Bu, C.B.C., C.C., T.F.

## Patient Authors

M.B. was involved at all stages of the research upholding patient perspective in design, and interpretation of results.

