## Supplement 1 for "Predicting long term clinical outcomes in Parkinson’s Disease using short term rating scales"

SUPPLEMENT 1: Additional detail concerning methods and results

1.0 **Methods**

Detailed definition of outcome measures

(i) Falls: A self-reported history of falls was ascertained at each follow-up by asking PwP “Do you fall?” having defined a fall as “…. an event which causes you to rest on the ground or other lower level unintentionally.” This was coded as: “no, never” [0], “yes, infrequently less than once per month” [1] and “yes, frequently more than once per month” [2]. We created two fall variables: “any falls” [1 or 2 versus 0] and “frequent falls” [2 versus 1 or 0] with the time of event associated with first occurrence on follow-up.

(ii) Dementia: We defined dementia as (a) Montreal Cognitive Assessment (MoCA) score of 21 or less and a score of 2 or more on the MDS-UPDRS item 1.1 (“cognitive impairment”) which provides supportive evidence that there is cognitive interference with normal activities and social interactions[1]. (b) Subjects classified with dementia at baseline had to have supportive evidence at the next visit to prevent misclassification from a random low in MoCA (“regression to the mean”). (c) If there was no confirmation then the individual was retained in the analysis and could potentially receive a subsequent classification of dementia. (d) If a subject was “withdrawn due to dementia”, we accepted the dementia classification even if there were missing data on MoCA and part I of the MDS-UPDRS.

(iii) Mortality: Deaths were ascertained either from next of kin contacting the study team or by the research team checking the Summary Care Record, a national database (SPINE portal) unless the patient opted out of sharing their data. Mortality data within the dataset represents all-cause mortality as cause of death was not available at the time of analysis.

**2.0 More detailed information about the results**

The dataset consisted of 3,667 observations from 958 participants (up until 17/05/2022) ranging from 1 (baseline only) to 8 (n=18) visits, with 77% having at least 4 visits and 33% at least 6 visits. The median age at first visit was 67.9 years (IQR: 61.4-74.1; range: 32.2-90.5), and Hoehn and Yahr stage 1=219 (23.0%), stage 2=659 (69.2%) and stage 3=73 (7.7%) and two at stage 4 and 5. 612 of 958 (63.9%) participants were male and the mean time since clinical diagnosis at baseline was 1.25 years (median=0.96; IQR=0.44-1.98; maximum=3.49 years). 956 had at least one UPDRS12 score measured at the first 3 visits and these were entered into the mixed model (934, 769 and 625 UPDRS12 scores at visits 1-3, respectively).

### **Modelling short-term disease progression using the combined UPDRS parts I and II**

The overall mean slope for *UPDRS12* was 2.08 (the mean increase per year; standard error [se]=0.11, median slope 1.96) with an intercept of 17.5 (se=0.33). The subject-specific slopes themselves had a standard deviation of 1.44 and a range of -3.24 to 8.96, IQR 1.12 to 2.81). Two subjects were excluded from all models as they had no UPDRS12 data in the first 3 years. Furthermore 42 patients had a slope less than 0 meaning that for the predictions involving the ‘treated’ cohort no change was made to their individual slope. Supplement 3 tabulates the mean slope for various subgroups based on death status and also number of missing visits. The mean slope for those who die is around 2.5 and those survive around 2.0, both before and after 3 years, suggesting missing visits in the model does not induce obvious bias in slope estimates.

### Dementia

The analysis dataset consisted of 121 events amongst 832 participants with 4,469 years at risk (median and maximum time at risk of 5.1 and 11.5 years, respectively). 18 subjects were excluded due to a baseline event and 106 excluded because they only had a baseline visit.

A unit increase in the *UPDRS12* slope was associated with an estimated hazard ratio (HR) of 1.52 (95% Confidence Interval [CI] 1.36-1.70; p<0.0001), indicating that an increase in the *UPDRS12* slope of 1 unit per year increases the hazard of *dementia* by 52%. Figure 1a) shows the marginal hazard functions (with 95% Cis) for the untreated (dark line) and treated (light line) assuming a 30% reduction. There is a large increase in the hazard over the first 3 years because of the exclusion of any cases at baseline. After 3-4 years, the hazard declines gently over the rest of the time-period but with widening uncertainty. The respective marginal survival predictions after a 10-year period were 72.6% and 78.4% (Figure 2a), meaning the cohort with the 30% lower trajectories might expect a 5.9% (95% CI: 4.3-7.4%) reduced risk of a *dementia* outcome (Figure 3a and Table 1a). This implies a number needed to treat to benefit (NNTB) of 16.9. Supplement 4 collates complete prediction results for all outcomes assuming a 30% and, as an addition, 50% reduction.

### Falls

For *any falls*, there were 396 events amongst 779 participants in 3,349 total years, where median and maximum time at risk was 3.6 and 11.5 years, respectively. 73 exclusions were due to a baseline event, 32 due to only data form a baseline visit and 72 due to no falls data at any visit.

The estimated HR was 1.37 (95% CI 1.29-1.46; p<0.0001), indicating that an increase in the *UPDRS12* slope of 1 unit per year increases the hazard of *any falls* by 37%. The shape of the (baseline) hazard function was similar to that of *dementia*, but with greater absolute values as this outcome is more common. The respective marginal survival probabilities at 10 years were much lower at 25.9% and 31.1% (Figure 2b), and the projected difference in survival probabilities was 5.2% (95%CI: 4.2-6.1% and NNTB=19.2) (Figure 3b and Table 1a).

There were only 88 *frequent falls* from 838 subjects with a median and maximum time at risk of 5.02 and 11.5 years, respectively. 43 were excluded for only having a baseline visit and 73 because they had no falls data from any visit.

The hazard ratio for *frequent falls* was larger in magnitude than for *any falls* (HR=1.68; 95%CI: 1.49-1.89; p<0.0001) though with greater uncertainty due to the smaller number of events. The hazard function was notably different to *any falls* and *dementia*, increasing monotonically, suggesting that once extant cases are removed, frequent falls was a milestone that increased in probability with PD progression (and age). 10-year marginal survival rates were 72.8% and 80.3% respectively (Figure 2c), with a larger survival difference of 7.5% (95%CI: 5.8-9.3% and NNTB=13.3) (Figure 3c and Table 1a).

### Mortality

There were 193 deaths in the 873 participants and 5,070 years at risk with a median and maximum time at risk of 6.0 and 11.5 years respectively. Baseline exclusions were restricted to those with only a baseline visit (n=83).

The *mortality* hazard ratio was 1.29 (95% CI 1.17-1.42; p<0.0001), around 30% higher for each additional unit increase of *UPDRS12.* The hazard function for *mortality* started from a very low base but rose and accelerated quickly, especially after year 6. Ten-year marginal survival probabilities were 46.6% and 50.6%, a survival difference of 4.0% (95%CI: 2.5-5.4% and NNTB=25.0) in favour of treatment (Figure 3d and Table 1).

1.	*The MoCA: Well-suited screen for cognitive impairment in Parkinson disease: Neurology: Vol 75, No 19.*
