## Supplementary figures and images for "Predicting long term clinical outcomes in Parkinson’s Disease using short term rating scales"

### Supplement 2

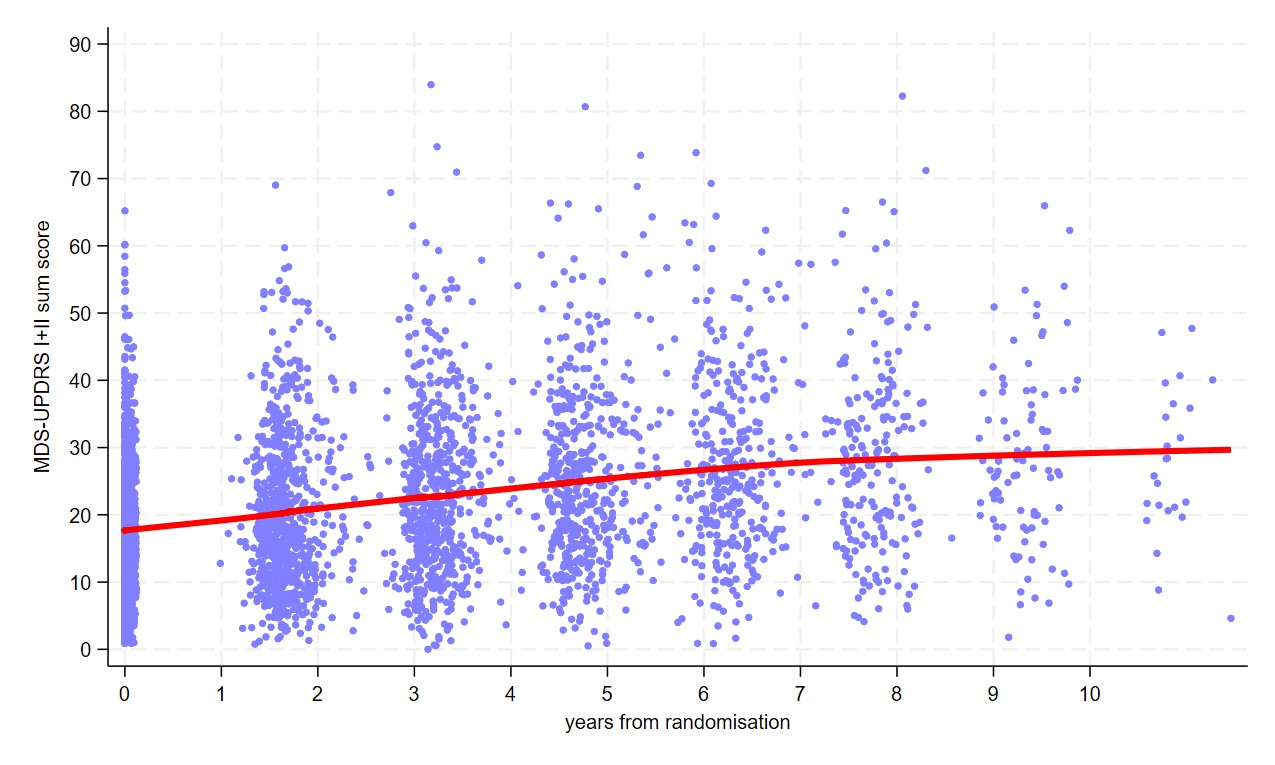

### Supplement 6

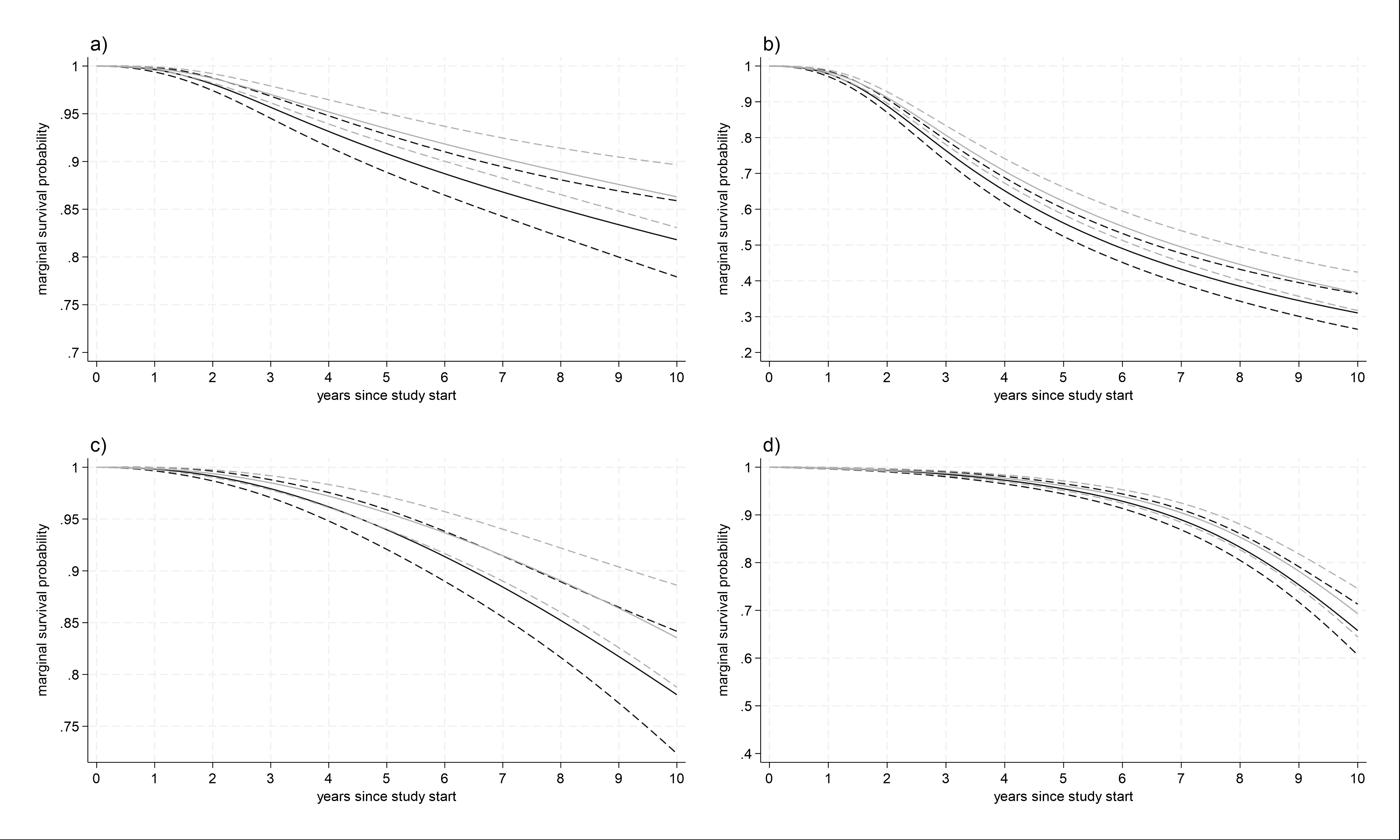

### Supplement 7

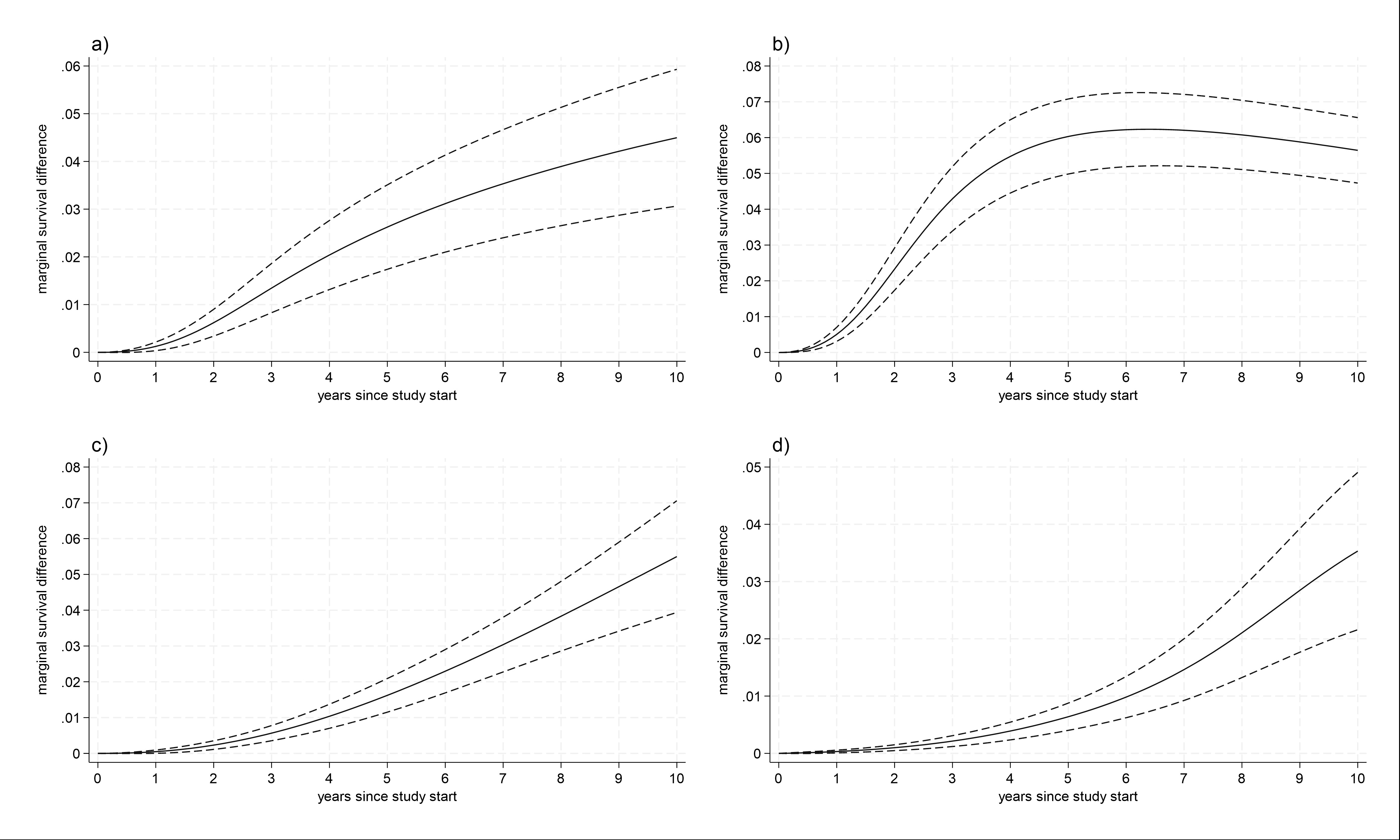
