## Supplement 3 for "Predicting long term clinical outcomes in Parkinson’s Disease using short term rating scales"

************************************************************
// Example code for creating individualised slopes for
// patients and fitting survival models to time to milestone
// and making marginalised survival predictions

// 
************************************************************


*** Individualised slopes 

/* need (with example names):

subject id variable: subjid - also dataset should be in 'long format' (type "help reshape" )
time variable for each outcome measure: time
shorter term clinical outcome variable measured at various time points including baseline: UPDRS12

also useful measure/visit label variable for easy inclusion/exclusion but could also use time variable or simply drop unwanted measures
*/


mixed UPDRS12 time  if visit<=3  || subjid: time, cov(unstr) // here only want to use for 3 visits (at 0, 1.5 and 3 years, though time is exact time)
predict ID_blup*, reffects relevel(subjid) // make random effect predictions for each patient: a random intercept (will be called ID_blup2) and a random slope(will be called ID_blup1)
gen ID_slope=ID_blup1+_b[time] // generate individual progression slope i.e. add random intercept to mean slope estimate

** then want to reshape to wide for survival part or simply drop repeated rows (consider "egen tag" or keep if visit==0 for example)

*** Survival model

** stset the dataset
/* need (with example names):

time to milestone  or censorship variable : milestone_time
indicator variable for if milestone (1)  or censorship (0): milestone

*/

stset milestone_time , failure(milestone==1) id(subjid) // id option in case there are multiple rows per ID

** flexible parametric survival model

/* need (with example names):

any covariates - think carefully if why including:
in our case 
age at BL - age
gender - gender
time since diagnosis to BL - disease_duration_diag

*/

stpm2 c.ID_slope c.age i.gender disease_duration_diag, df(2) scale(h)  eform lininit // hazard scale preferable if wish hazard ratios. Use likelihood ratio tests (or AIC) to determine appropriate df but also look a implied hazard plots to see if makes sense and have not overfitted

stpm2 c.ID_slope##c.age i.gender disease_duration_diag, df(2) scale(h)  eform lininit // an example with an interaction term too (slope#age interaction here)

*** predictions

** first create 'treatment' cohort whose progression has been improved by 30%
gen ID_slope_30pct=ID_slope
replace ID_slope_30pct=ID_slope*0.70 if ID3_slope>=0 (those with progression <=0 left 'unimproved')
range timevar 0 10 101 // create time variable for predictions

 ** hazard predictions for actual sample plus 'treatment cohort'extra preds at 30%

standsurv if e(sample),hazard atvars(haz_control haz_trt30)  at1(.) at2(ID_slope= ID_slope_30pct ) timevar(timevar) ci 

 ** survival predictions for actual sample plus 'treatment cohort'extra preds at 30%

standsurv if e(sample), atvars(S_control S_trt30)  at1(.) at2(ID_slope= ID_slope_30pct ) timevar(timevar) ci 

 ** survival difference predictions between actual sample and 'treatment cohort'extra preds at 30%

standsurv if e(sample), atvars(at1 at2 ) at1(. ) at2(ID_slope= ID_slope_30pct) timevar(timevar) ci   contrast(difference) contrastvar(Sdiff30)
drop at*

 ** additional survival difference predictions for 'younger' cohort
 
 qui su age
 local agediff60=r(mean)-60
 gen age_mean60=age-`agediff60' // create younger cohort, here universally made younger by amount so that new mean age=60 

standsurv if e(sample), atvars(at1 at2 ) at1(age=age_mean60 ) at2(ID2_slope= ID2_slope_30pct age=age_mean60  ) timevar(timevar) ci   contrast(difference) contrastvar(Sdiff30_ageM60)
drop at*
