## Supplement 8 for "Predicting long term clinical outcomes in Parkinson’s Disease using short term rating scales"

**Supplementary Material. EJS ACT-PD Consortium (20th February 2024)**

| **Member** | **Role** | **Organisation** | **Email** |
| --- | --- | --- | --- |
| Thomas Foltynie | Co-Lead | University College London, London, UK | |
| Camille B Carroll | Co-Lead | Newcastle University, Newcastle, UK;  University of Plymouth, Plymouth, UK | |
| Roger Barker | Chair | University of Cambridge, Cambridge, UK | |
| James Carpenter | Chair | MRC Clinical Trials Unit at UCL, London, UK |.ac.uk |
| Yoav Ben Shlomo | Member | University of Bristol, Bristol, UK |.ac.uk |
| Mark Edwards | Member | St George’s University of London, London, UK | |
| Alan Whone | Member | University of Bristol, Bristol, UK |.ac.uk |
| Carl Counsell | Member | University of Bristol, Bristol, UK |.ac.uk |
| Caroline S Clarke | Member | University College London, London, UK |.ac.uk |
| Matthew Burnell | Member | MRC Clinical Trials Unit at UCL, London, UK |.ac.uk |
| Kate Hockey | PPI | Expert by Experience | |
| Anna Jewell | PPI | Expert by Experience |.com |
| Priti Gros | Member | University of Toronto, Toronto, Canada |.ca |
| Tom Barber | Early Career Researcher (ECR) | University of Oxford, Oxford, UK | |
| Anette Schrag | Chair | University College London, London, UK |.ac.uk |
| Rimona S Weil | Deputy Chair | University College London, London, UK |.ac.uk |
| Caroline H Williams-Gray | Member | University of Cambridge, UK | |
| Michele T Hu | Member | University of Oxford, Oxford, UK |.ox.ac.uk |
| Lynn Rochester | Member | Newcastle University, Newcastle, UK |.ac.uk |
| Paola Piccini | Member | Imperial College London, London UK |.ac.uk |
| Henrik Zetterberg | Member | University College London, London, UK / University of Gothenburg, Mölndal, Sweden | |
| Alastair Noyce | Member | Queen Mary University of London, London UK | |
| Michael Lawton | Member | University of Bristol, Bristol, UK | |
| Ashwani Jha | Member | University College London, London, UK | |
| Brook Huxford | Member | Queen Mary University of London, London, UK | |
| Shlomi Haar Millo | Member | Imperial College London, London, UK | |
| K. Ray Chaudhuri | Member | King's College London, London, UK | |
| Carroll Siu | PPI | Expert by Experience | |
| Michèle Bartlett | PPI | Expert by Experience | |
| Kuhan Pushparatnam | PPI | Expert by Experience | |
| Daniel van Wamelen | ECR | King's College London, London, UK | |
| Anthony HV Schapira | Co-Chair | University College London, London, UK | |
| Oliver Bandmann | Co-Chair | University of Sheffield, Sheffield, UK | |
| Simon Stott | Member | Cure Parkinson's, London, UK | |
| George Tofaris | Member | University of Oxford, Oxford, UK | |
| Esther Sammler | Member | University of Dundee, Dundee, UK | |
| Heather Mortiboys | Member | University of Sheffield, Sheffield, UK | |
| Li Wei | Member | University College London, London, UK | |
| Alan Wong | Member | Royal Free Hospital NHS Foundation Trust, London, UK | |
| Susan Duty | Member | King's College London, London, UK | |
| David Dexter | Member | Parkinson's UK, London, UK | |
| Keith Martin | PPI | Expert by Experience | |
| Edwin Jabbari | ECR | University College London, London, UK | |
| Stephen Mullin | Chair | University of Plymouth, Plymouth, UK | |
| Huw Morris | Member | University College London, London, UK | |
| David Breen | Member | University of Edinburgh, Edinburgh, UK | |
| Christian Lambert | Member | University College London, London, UK | |
| Prasad Korlipara | Member | University College London, London, UK | |
| Monty Silverdale | Member | University of Manchester, Manchester, UK | |
| Kailash Bhatia | Member | University College London, London, UK | |
| Alison Yarnall | Member | Newcastle University, Newcastle, UK | |
| Raj Khengar | Member | University College London, London, UK | |
| Helen Collins | Member | Nat National Institute of Health Research Clinical Research Network, UK | |
| Fleur Hudson | Member | MRC Clinical Trials Unit at UCL, London, UK. | |
| Rebecca Croucher | Member | National Institute of Health Research Clinical Research Network, UK | |
| Sandra Bartolomeu-Pires | Member | Southhampton NHS Foundation Trust, Southampton, UK | |
| Veena Agarwal | Member | Southhampton NHS Foundation Trust, Southampton, UK | |
| Jennifer Allison | Member | National Institute of Health Research Clinical Research Network, UK | |
| Jodie Forbes | PPI | Expert by Experience | |
| Alex Edwards | Member | Parkinson’s UK, London, UK | |
| Sheila Wonnacott | PPI | Expert by Experience | |
| Dilan Athauda | ECR | University College London, London, UK | |
| Joy Duffen | Co-Chair | Cure Parkinson’s, London, UK | |
| Sonia Gandhi | Co-Chair | University College London, London, UK | |
| Emily Henderson | Member | University of Bristol, Bristol, UK | |
| Jen Black | Member | University College London, London, UK | |
| Shona Clegg | Member | Parkinson's UK, London, UK | |
| Karen Matthews | Member | National Institute of Health Research Clinical Research Network, UK | |
| Vince Greaves | Member | University College London, London, UK | |
| Eric Deeson | PPI | Expert by Experience | |
| Laurel Miller | PPI | Expert by Experience | |
| Joel Handley | ECR | Salford Royal NHS Foundation Trust, UK | |
| Sarah Medd Phillips | Member | University College London, London, UK | |
| Helen Matthews | Member | Cure Parkinson’s, London, UK | |
| Kevin McFarthing | Chair | Expert by Experience | |
| Amit Batla | Member | University College London, London, UK | |
| Nikul Bakshi | Member | Parkinson's UK, London, UK | |
| Emma Lane | Member | UCB Pharma | |
| Miriam Parry | Member | Kings College Hospital NHS Foundation Trust, London, UK | |
| Natasha Ratcliffe | Member | Independent Advisor, Manchester, UK | |
| Cheney Drew | Member | Cardiff University, Cardiff, UK | |
| Naveena Kapur | Member | Parkinson's UK, London, UK | |
| Anaya Navangul | Member | Cure Parkinson's, London, UK | |
| Shafaq Ali | Member | Expert by Experience | |
| Katherine Fletcher | Member | Parkinson's UK, London, UK | |
| Claire Bale | Member | Parkinson's UK, London, UK | |
| Cristina Gonzalez-Robles | Member | University College London, London, UK | |
| Marie-Louise Zeissler | Member | University of Plymouth, Plymouth, UK | |
| Georgia Mills | Member | University College London, London, UK | |
| Romy Ellis-Doyle | Member | University College London, London, UK | |
| Sally Collins | Member | University of Plymouth, Plymouth, UK | |
| Rebecca Petty | Member | University of Plymouth, Plymouth, UK | |

Abbreviations: PPI - Patient and Public Involvement Representative
